# Fibrinolysis resistance in sepsis assessed by serial beside viscoelastometry is associated with plasma protein profiles and organ-failure severity: a prospective observational study

**DOI:** 10.64898/2026.10.24.26366401

**Authors:** Lucy Coupland, Matthew Self, Eurwin Suryana, Ruby Law, Philip Crispin, Robert Medcalf, Charithani Keragala, Steve Frost, David Rabbolini, Peter K Moore, Anders Aneman

## Abstract

**Background:** Fibrinolysis resistance is associated with adverse outcomes in sepsis and can be quantified at the bedside by tissue plasminogen activator–enhanced viscoelastometric testing, using the fibrin-adjusted (FIBA10) clot lysis time (TPA-LT) expressed as the TPA-LT/FIBA10 ratio. In this study of adults admitted to intensive care with sepsis, plasma tissue plasminogen activator activity, plasminogen activator inhibitor-1 activity (PAI-1), plasminogen and α2-antiplasmin were measured serially alongside TPA-LT/FIBA10 and Sequential Organ Failure Assessment (SOFA-2) scores. Whether the bedside TPA-LT/FIBA10 measure of fibrinolysis resistance reflects the underlying plasma fibrinolytic protein environment, and whether it tracks organ-failure severity, has not been established.

**Results:** This prospective observational study included 283 paired observations in 69 septic patients admitted with a median Acute Physiology and Chronic Health Evaluation III score 81, median SOFA-2 score 13 and a 38% ICU mortality. PAI-1 increased and plasminogen decreased with increasing fibrinolysis resistance (1.77 [1.50-2.09]-fold increase and 0.81 [0.76-0.87]-fold decrease, respectively, per e-fold increase in TPA-LT/FIBA10). Increasing organ-failure severity was also associated with increased PAI-1 and decreased plasminogen (1.23 [1.17–1.29]-fold increase and 0.95 [0.93–0.96]-fold decrease, respectively, per point increase in SOFA-2). In contrast, tissue plasminogen activator activity and absolute α2-antiplasmin did not relate to fibrinolysis resistance or organ failure. Principal component analysis of the fibrinolysis proteins, organ-failure severity and fibrinolysis resistance identified a dominant axis explaining 35% of the variance, on which PAI-1, SOFA-2 and TPA-LT/FIBA10 loaded together in opposition to plasminogen. Unsupervised clustering of the dataset, performed without reference to patient outcomes, separated a fibrinolysis inhibitor-high, substrate-depleted group with 61% mortality compared to 29% in the preserved-fibrinolysis group. During the ICU stay, PAI-1 decreased within 72 hours in survivors but remained unchanged in non-survivors (interaction p<0.001). A trend was observed for plasminogen levels increasing in survivors but remaining static in non-survivors.

**Conclusions:** Changes in key fibrinolysis proteins, organ-failure severity and fibrinolysis resistance measured by viscoelastometry were coupled facets of the response to sepsis dominated by fibrinolysis inhibitor excess and substrate depletion rather than by reduced activator availability. These findings provide biological validation for bedside viscoelastometric quantification of fibrinolysis resistance and support prospective evaluation of substrate-directed profibrinolytic strategies.

**ESSENTIALS:**

- Fibrinolysis resistance (FR) in sepsis associates with organ failure and poor outcomes
- Repeat FR assays in 69 septic ICU cases using viscoelastometry, clinical and protein evaluation
- FR linked to organ failure via joint decrease in fibrinolysis substrate and increased inhibition
- Grading fibrinolysis resistance by viscoelastometry in sepsis quantifies clinical risk

## BACKGROUND

Fibrinolysis resistance is increasingly recognised as a clinically important phenotype associated with adverse outcomes in critically ill patients admitted to the intensive care unit (ICU) with sepsis [1–3]. A grading system was recently proposed that enables the degree and dynamics of fibrinolysis resistance to be ascertained at the point-of-care using viscoelastometric testing (VET) and which was associated with ICU outcomes [4, 5]. The variable underlying the grade, obtained using a ClotPro^®^ device, is the clot lysis time in the tissue plasminogen activator (tPA)-enhanced assay (TPA-LT, seconds), normalised to functional fibrinogen as measured by the fibrin-specific clot amplitude at 10 minutes (FIBA10, mm), and expressed as TPA-LT/FIBA10 (seconds/mm).

Sepsis leads to excessive thrombin generation [6] and deposition of dense fibrin networks which are reported to assist pathogen sequestration [7, 8]. At the same time, fibrinolysis is reduced by increased production of the principal physiological inhibitor of fibrinolysis, plasminogen activator inhibitor type 1 (PAI-1), and the primary inhibitor of plasmin, α2-antiplasmin (α2AP) [3, 9, 10]. The role of thrombin-activatable fibrinolysis inhibitor (TAFI) remains equivocal [1] with studies reporting both positive [11] and negative [12] associations. Levels of the key fibrinolysis substrate plasminogen [13] may become depleted and represent another mechanism contributing to fibrinolysis resistance in sepsis [14, 15] Tissue plasminogen activator levels typically show a biphasic response in sepsis, with an early transient rise that is rapidly overwhelmed by sustained PAI-1 elevation, such that the fibrinolytic state in the established ICU phase may be determined more by inhibitor excess and substrate availability than by activator supply [1]. Overall, these changes result in a fibrinolysis resistant state that exacerbates the pro-thrombotic response associated with sepsis.

Since plasma protein levels cannot be determined within a clinically actionable timeframe in the routine clinical setting, it is important to establish how the VET-derived TPA-LT/FIBA10 quantification of fibrinolysis resistance is reflected in changes in plasma levels of key fibrinolysis proteins. Understanding the associations between TPA-LT/FIBA10, changes in key fibrinolysis proteins and organ failure severity is important to determine the biological validity of this bedside test. Furthermore, insights into the biological mechanism(s) behind fibrinolysis resistance are critical to any therapeutic interventions.

In this prospective observational cohort study of ICU patients with sepsis, plasma fibrinolysis proteins (tPA activity, PAI-1 activity, plasminogen and α2AP concentrations) were measured serially, alongside contemporaneous TPA-LT/FIBA10 measures and Sequential Organ Failure Assessment (SOFA-2) scores [16]. The primary aim was to characterise the plasma protein basis of the TPA-LT/FIBA10 variable and its relationship to organ-failure severity, with ICU mortality examined as a secondary, exploratory outcome. It was hypothesised that increased TPA-LT/FIBA10 would be accompanied by a coherent plasma protein signature of inhibitor excess and substrate depletion and that this signature would track organ-failure severity.

## METHODS

This is a substudy of the FORTITUDE single-centre, prospective observational study in sepsis (NCT06680180) conducted at Liverpool ICU, Australia, between January 2023 and April 2026. The study was approved by the South Western Sydney Local Health District Human Research Ethics Committee (2022/ETH02122) with a waiver of consent as permitted under the National Statement on Ethical Conduct in Human Research 2023 for low-risk research in critically ill patients. The study adheres to the STROBE guidelines for observational studies (see Table S1, Additional file 1).

### Participants

Adult patients admitted to the ICU with a diagnosis of sepsis by Sepsis-3 criteria [17], without any limitations of medical therapy and expected to survive beyond the day following admission were screened for inclusion. Exclusion criteria were pre-existing chronic coagulation disorder, known pregnancy, or admission with palliative intent. Staff trained in performing VET needed to be available with the first VET performed within 24 hours of admission. All VET data were collected prospectively with anonymised clinical and pathology data added retrospectively including the results of protein analyses.

### Clinical data

Baseline data included patient characteristics, APACHE III (Acute Physiology and Chronic Health Evaluation), SIC (Sepsis Induced Coagulopathy) [18], DIC (disseminated intravascular coagulation) [19] and CFS (Clinical Frailty Score) [20] scores. All clinical data, blood test results and support measures needed to calculate the SOFA-2 score [16] were sourced from the electronic medical records system. In addition, the use of therapeutic steroids and anticoagulants was captured.

### Viscoelastometric assays and fibrinolysis assessment

All VET was performed using the ClotPro® (Haemoview Diagnostics, Milton, QLD, Australia) platform within 30 minutes of each blood draw as previously described [4, 5]. In short, blood was collected into 3.2% buffered sodium citrate and three assays conducted: (i) EX-test - whole blood coagulation activated with tissue factor with a heparin inhibitor included; (ii) FIB-test - same as the EX-test with two platelet inhibitors included to assess fibrin clot formation; and (iii) TPA-test - same as the EX-test with tPA (650 ng/mL) added to permit evaluation of fibrinolysis. The following VET results were documented: the time for the forming clot to reach 2 mm amplitude in the EX-test (EX-CT, sec); clot amplitudes 10 min after commencement of clot formation in the FIB- and EX-tests (FIBA10 and EXA10, mm); the time taken for 50% of the maximum clot firmness to be lysed in the TPA-test (TPA-LT, sec); and maximum clot firmness/amplitude measured during the 60-minute assay run time in the TPA test (TPA MCF, mm). A fibrin-adjusted measure of fibrinolysis was obtained by dividing the TPA-LT by the FIBA10 (TPA-LT/FIBA10, sec/mm) [4]. An ordinal grading system, as previously described, based on the continuous TPA-LT/FIBA10 was used to classify increasing fibrinolysis resistance: Grade 1 if TPA-LT/FIBA10 <13 sec/mm, Grade 2 if TPA-LT/FIBA10 ≥13 but <26 and Grade 3 if TPA-LT/FIBA10 ≥26 [4]. All VET assays were performed in patients remaining in ICU on days 1, 2, 3, 5, 7, 10 and 15 after admission, with the timepoints referred to as episodes. The following terminology is used throughout the manuscript ‘TPA-LT/FIBA10’ when referring to the viscoelastic testing measurement of fibrinolysis resistance; ‘resistance grade’ when referring to the TPA-LT/FIBA10 derived grading system; and ‘fibrinolysis resistance’ when referring to the biological state.

### Measurements of plasma fibrinolysis proteins

Contemporaneous to each VET assay, blood in 3.2% buffered sodium citrate was centrifuged at 5000 x g for 20 min at room temperature to obtain platelet-poor plasma that was frozen at − 80°C. ELISA kits (Innovative Research, Novi, MI, USA) were used to measure tPA activity (IU/mL) (IHUTPAKT), PAI-1 activity (IU/mL) (IHUPAI1KT), plasminogen concentration (µg/mL) (IHUPLGKT) and α2AP concentration (µg/mL) (IHUA2APLKTT). Plasma samples were diluted and processed following the manufacturer’s instructions. Staff performing the ELISA were blinded to clinical and VET data.

### Statistical methods

No formal power calculation was performed for this exploratory study. Patient characteristics, ICU interventions, VET variables and fibrinolysis proteins are reported as medians [interquartile range, IQR] as appropriate for non-normally distributed data or as counts (%). The primary outcome was organ failure defined by the SOFA-2 score, analysed both continuously and in tertiles. Survival in ICU was used as a secondary outcome since the study was not powered for mortality. All fibrinolysis protein data showed a right-skewed distribution and were therefore log transformed in the statistical modelling. Differences in fibrinolysis protein concentrations split by fibrinolysis resistance or SOFA-2 tertile were analysed by a linear mixed-effects model, with resistance or tertile set as a fixed effect with a random intercept for patient to account for within-subject correlation. Associations are report per e-fold (∼2.72x) increase in fibrinolysis resistance and per one point increase in SOFA-2. The relationship between each fibrinolysis protein and the continuous log(TPA-LT/FIBA10) are shown with a log cubic spline fit with 95% confidence intervals (95%CI) from 1000 bootstraps and a linear log-log fit. The Spearman correlation and the fold change in protein per doubling (2x) of the log(TPA-LT/FIBA10) are reported.

To characterise the joint structure of the fibrinolysis and organ failure severity variables, principal component analysis (PCA) was performed on the six standardised variables: log(tPA), log(PAI-1), log(plasminogen), log(α2AP), log(TPA-LT/FIBA10) and SOFA-2 score. Ward’s minimum-variance hierarchical clustering (k=2) was applied to the same standardised matrix using Euclidean distance, and the resulting unsupervised clusters were described and related post hoc to ICU outcome. The number of clusters was explicitly set to two *a priori* to facilitate interpretability (e.g. a clinical interpretation of high-risk vs. low-risk). Clusters are shown with the patient level centroid ellipse and its 95%CI, and the data ellipse for individual observations to show data concentration including the 95%CI.

The trajectories for fibrinolysis proteins across sampling episodes in ICU are shown by the mixed-effects model with LOESS smoothing. Differences were explored using an episode-by-organ failure severity or ICU outcome interaction on log-fibrinolysis protein. Divergence between groups was assessed by a likelihood-ratio test comparing the interaction model with an additive (parallel-trajectory) model fitted by maximum likelihood, supported by the Akaike information criterion. Statistical inference was restricted to episodes 1–3, for which group sizes were adequate.

The main statistical analyses were performed using observational level data treating each episode as an independent event. In sensitivity analyses, all data were also modelled at the patient level with between and within differences assessed separately (see Figures S4-S6, Additional file 1).

Statistical significance was set at a two-sided p-value of <0.05. No corrections for multiple testing were performed in this exploratory study, and the results are considered hypothesis-generating. All analyses were performed in RStudio (Integrated Development Environment for R, version 2023.06.0, Posit Software, PBC, Boston MA, USA) using R 4.3.3 (R Core Team, 2024). The R scripts were coded with assistance from Claude (Sonnet 4.6, Anthropic 2025, https://www.anthropic.com). All code, outputs and interpretations were reviewed and verified by the authors.

## RESULTS

The study included 69 patients with 283 complete sets of VET assays and fibrinolysis proteins obtained for 4 [1–7] episodes. Data were collected at 23 [IQR 16–25] hour intervals for the first 3 episodes, at 48 [45–50] hour intervals for episodes 5 and 7, and 120 hour interval for episodes 10 and 15, with few patients remaining in ICU at these time points (see Figure S1, Additional file 1). The patient characteristics on admission to ICU are reported in Table 1. The cohort was severely ill with a median APACHE III score of 81 and median baseline SOFA-2 score of 13. Twenty-six patients (38%) died in ICU. The ICU mortality increased across baseline SOFA-2 tertiles (12%, 33% and 57% in the low, middle and high tertiles) similar to that observed with increasing fibrinolysis resistance (19%, 35% and 54% for Grades 1–3).

**Table 1.** Patient characteristics, organ support and coagulation variables split by fibrinolysis resistance grade, SOFA-2 tertile and ICU outcome.

|  | Fibrinolysis grade |  |  |  | SOFA-2 tertile |  |  | ICU outcome |  |
| --- | --- | --- | --- | --- | --- | --- | --- | --- | --- |
|  | All<br>(n=69) | Grade 1<br>(n=21) | Grade 2<br>(n=20) | Grade 3<br>(n=28) | ≤10<br>(n=18) | 11–13<br>(n=21) | ≥14<br>(n=30) | Survived<br>(n=43) | Died<br>(n=26) |
| **Admission demographics and illness severity** |  |  |  |  |  |  |  |  |  |
| Age<br>(years) | 67<br>[49–77] | 76<br>[67–78] | 76<br>[46–81] | 57<br>[50–67] | 76<br>[68–77] | 67<br>[51–78] | 56<br>[45–74] | 68<br>[48–76] | 63<br>[50–79] |
| Male<br>(n, %) | 34 (49) | 8 (38) | 13 (65) | 13 (46) | 4 (22) | 12 (57) | 18 (60) | 14 (33) | 20 (77) |
| CFS | 4 [3–6] | 4 [3–5] | 4 [3–6] | 4 [3–4] | 4 [3–5] | 6 [3–6] | 4 [3–4] | 4 [3–6] | 3 [2–6] |
| APACHE III | 81<br>[68–103] | 62<br>[57–70] | 85<br>[74–101] | 86<br>[74–113] | 80<br>[62–87] | 73<br>[67–107] | 83<br>[71–104] | 73<br>[64–88] | 98<br>[74–137] |
| SOFA-2 | 13<br>[11–14] | 12<br>[10–14] | 13<br>[10–14] | 14<br>[11–15] | 9<br>[8–10] | 12<br>[11–13] | 14<br>[14–15] | 12<br>[10–14] | 14<br>[12–15] |
| Lactate<br>(mmol/L) | 2.0<br>[1.6–2.7] | 1.7<br>[1.6–2.0] | 2.0<br>[1.3–2.6] | 2.4<br>[2.0–6.8] | 2.1<br>[1.8–2.8] | 1.7<br>[1.2–2.6] | 2.1<br>[1.8–2.6] | 1.8<br>[1.4–2.4] | 2.5<br>[1.9–5.7] |
| **Organ support and length of stay** |  |  |  |  |  |  |  |  |  |
| VIS | 25<br>[12–43] | 13<br>[8.2–14] | 42<br>[11–47] | 30<br>[25–91] | 12<br>[9.5–20] | 24<br>[11–32] | 34<br>[25–68] | 25<br>[11–42] | 26<br>[13–72] |
| Ventilation<br>(n, %) | 56 (81) | 13 (62) | 15 (75) | 28 (100) | 12 (67) | 18 (86) | 26 (87) | 37 (86) | 19 (73) |
| Dialysis<br>(n, %) | 28 (41) | 4 (19) | 6 (30) | 18 (64) | 5 (28) | 9 (43) | 14 (47) | 14 (33) | 14 (54) |
| Steroid therapy<br>(n, %) | 37 (54) | 6 (29) | 10 (50) | 21 (75) | 4 (22) | 8 (38) | 25 (83) | 18 (42) | 19 (73) |
| Anticoagulation<br>(n, %) | 5 (9) | 3 (12) | 2 (12) | 0 (0) | 2 (8) | 3 (25) | 0 (0) | 4 (9) | 1 (4) |
| ICU LOS<br>(days) | 6.7<br>[2.7–12] | 6.0<br>[1.7–11] | 8.8<br>[5.6–11] | 6.7<br>[3.6–13] | 3.8<br>[1.7–7.1] | 8.9<br>[4.2–13] | 7.4<br>[2.5–13] | 7.3<br>[3.6–11] | 6.7<br>[2.5–12] |
| Hospital LOS<br>(days) | 16<br>[8.8–28] | 16<br>[9.8–23] | 15<br>[5.3–32] | 18<br>[12–61] | 16<br>[7.1–22] | 16<br>[12–20] | 33<br>[12–64] | 17<br>[9.7–32] | 7.1<br>[5.1–11] |
| **Coagulation on admission** |  |  |  |  |  |  |  |  |  |
| SIC score | 4<br>[2–4] | 3<br>[2–4] | 3<br>[3–4] | 4<br>[3–4] | 3<br>[2–4] | 3<br>[2–4] | 4<br>[3–5] | 3<br>[2–4] | 4<br>[3–5] |
| DIC score | 4<br>[3–5] | 3<br>[3–4] | 4<br>[2–4] | 4<br>[3–5] | 3<br>[2–4] | 3<br>[2–4] | 4<br>[3–5] | 3<br>[3–4] | 4<br>[2–5] |
| PT<br>(sec) | 14<br>[13–17] | 14<br>[13–16] | 14<br>[13–17] | 16<br>[13–24] | 14<br>[13–17] | 14<br>[13–16] | 15<br>[13–19] | 14<br>[13–16] | 15<br>[13–23] |
| INR | 1.3<br>[1.2–1.6] | 1.3<br>[1.2–1.4] | 1.2<br>[1.2–1.6] | 1.5<br>[1.2–2.2] | 1.2<br>[1.2–1.6] | 1.2<br>[1.2–1.5] | 1.4<br>[1.2–1.7] | 1.2<br>[1.2–1.5] | 1.4<br>[1.2–2.1] |
| Platelets<br>($\times 10^9/L$) | 183<br>[100–246] | 232<br>[140–295] | 141<br>[56–176] | 215<br>[144–244] | 180<br>[137–203] | 238<br>[147–313] | 147<br>[51–244] | 215<br>[141–282] | 147<br>[59–222] |
| D-dimers<br>(mg/L FEU) | 5.6<br>[2.9–12] | 5.6<br>[2.2–16] | 5.2<br>[2.7–13] | 5.0<br>[3.2–6.6] | 5.0<br>[3.5–6.0] | 6.3<br>[2.8–13] | 3.8<br>[3.1–15] | 5.6<br>[2.8–9.4] | 5.1<br>[3.1–17] |
| **Viscoelastometry (ClotPro) on admission** |  |  |  |  |  |  |  |  |  |
| EX CT<br>(sec) | 75<br>[62–88] | 68<br>[63–78] | 75<br>[60–83] | 87<br>[73–97] | 70<br>[68–83] | 63<br>[56–86] | 83<br>[65–88] | 70<br>[61–85] | 79<br>[64–92] |
| EX A10<br>(mm) | 61<br>[57–65] | 63<br>[60–64] | 58<br>[53–60] | 65<br>[52–69] | 60<br>[60–63] | 63<br>[61–69] | 53<br>[50–63] | 63<br>[61–67] | 57<br>[52–61] |
| FIB CT | 90 | 87 | 76 | 109 | 85 | 72 | 93 | 87 | 98 |
| (sec) | [74–110] | [64–98] | [73–92] | [91–122] | [76–99] | [67–120] | [90–110] | [75–106] | [73–111] |
| FIB A10 | 25 | 25 | 23 | 31 | 25 | 27 | 25 | 27 | 23 |
| (mm) | [21–32] | [25–26] | [20–28] | [20–34] | [24–27] | [23–35] | [12–31] | [25–33] | [14–26] |
| TPA MCF | 41 | 37 | 36 | 61 | 37 | 49 | 40 | 41 | 40 |
| (mm) | [36–54] | [37–41] | [28–41] | [48–63] | [37–43] | [41–63] | [29–49] | [37–60] | [29–49] |
| TPA LT | 480 | 242 | 420 | 2730 | 358 | 420 | 615 | 420 | 615 |
| (sec) | [262–1150] | [235–276] | [295–510] | [1045–3600] | [293–621] | [240–3600] | [260–1150] | [263–1150] | [269–1301] |
| TPA-LT/FIB A10 | 17.5 | 10.0 | 16.2 | 85.7 | 16.8 | 13.8 | 26.7 | 15.6 | 26.7 |
|  | [11.4–49.4] | [9.4–11.3] | [14.5–20.9] | [37.1–109.1] | [11.3–29.3] | [11.1–102.9] | [13.8–78.8] | [11.2–33.9] | [15.0–102.9] |
| **Outcome** |  |  |  |  |  |  |  |  |  |
| ICU death<br>(n, %) | 26 (38) | 4 (19) | 7 (35) | 15 (54) | 2 (12) | 7 (33) | 17 (57) | 0 (0) | 26 (100) |

Data are median [interquartile range] or count (n) and percentage (%) as assessed on admission to ICU, corresponding to the first sampling episode.
Steroid therapy was defined as >200 mg hydrocortisone *or* >8 mg dexamethasone *or* >500 mg methylprednisolone daily.
Anticoagulation was defined as therapeutic doses of >10,000 units unfractionated heparin *or* >40 mg enoxaparin daily.
CFS, Clinical Frailty Scale; APACHE, Acute Physiology and Chronic Health Evaluation; SOFA, Sequential Organ Failure Assessment; VIS, Vasopressor Inotrope Score; LOS, Length of stay; SIC, Sepsis Induced Coagulopathy; DIC, Disseminated Intravascular Coagulation; PT, prothrombin time; INR, international normalised ratio; EX/FIB/TPA, ClotPro assays; CT, clotting time; A10, amplitude at 10 min; MCF, maximum clot firmness; LT, lysis time.

### Plasma fibrinolysis proteins in relation to organ failure severity and fibrinolysis resistance

The correlations between fibrinolysis protein levels and continuous TPA-LT/FIBA10 are shown in Figure 1. The strongest association was found for PAI-1 (Spearman ρ = +0.54, p<0.001, 1.62-fold increase), with plasminogen inversely related (ρ = −0.26, p<0.001, 0.86-fold decrease). Neither tPA activity (ρ = −0.02) nor α2AP (ρ = −0.08) were statistically significantly related to fibrinolysis resistance. Plasma analyses of fibrinolysis proteins are reported in Table 2 (and in Additional file 1, Figures S2A and S2B) split by fibrinolysis resistance grades, SOFA-2 tertiles and ICU outcome. The levels of PAI-1 increased with fibrinolysis resistance grade (median 14, 35 and 131 IU/mL for Grades 1–3), across SOFA-2 tertiles (17, 27 and 117 IU/mL, respectively) and in ICU non-survivors. In contrast, plasminogen had an inverse relationship to fibrinolysis grade (median 131, 104 and 77 µg/mL for Grades 1–3) and increasing SOFA-2 score tertiles (median 108, 109 and 79 µg/mL). The linear-trend was significant (p<0.001) for each variable. Median PAI-1 was 9.4-fold higher and plasminogen 41% lower in Grade 3 fibrinolysis resistance compared to Grade 1. The activity of tPA and α2AP concentration showed no significant differences across SOFA-2 tertiles or fibrinolysis resistance grades. These findings indicate that the bedside fibrinolysis resistance grade stratified an inhibitor-driven and substrate-depleted state of reduced fibrinolysis rather than differences in tPA availability. Thus organ-failure severity and fibrinolysis resistance grade obtained using bedside VET mirror a shared, progressively worsening state of impaired fibrinolysis in sepsis. The associations between plasma fibrinolysis protein levels and increased fibrinolysis resistance (per e-fold, ∼ 2.72-fold) and SOFA-2 score (per point) are reported in Table 3.

**Figure 1.**
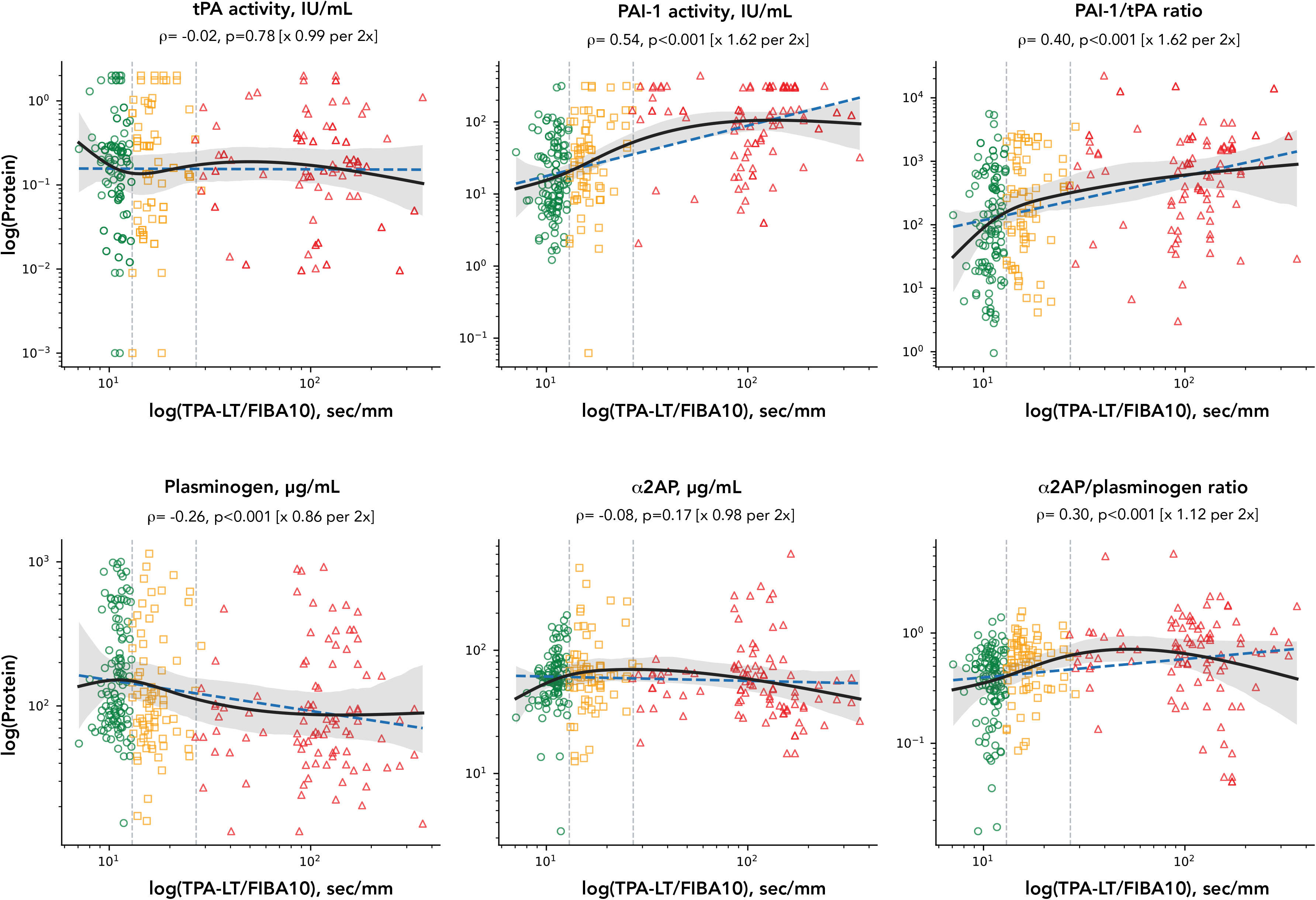
Plasma fibrinolysis proteins in relation to fibrinolysis resistance measured bedside by tPA-enhanced VET. Each fibrinolysis protein (log scale) is plotted against the concurrent fibrin-adjusted lysis time, TPA-LT/FIBA10 (log scale, sec/mm), across all observations. The vertical dashed lines represent the boundaries between fibrinolysis resistance grades: Grade 1 (<13 sec/mm), green circles; Grade 2 (≥13 to <26 sec/mm), amber squares; Grade 3 (≥26 sec/mm), red triangles. The solid black line represents the natural cubic spline fit with its 95% confidence interval. The dashed blue line represents the linear log–log fit. No spline fit was significantly different from the line fit across proteins supporting linear relations to TPA-LT/FIB10. Each panel reports the Spearman correlation coefficient (ρ) with its p value and the fold-change in protein per doubling (2x) of TPA-LT/FIBA10 in brackets.

**Table 2.** Plasma fibrinolysis protein concentrations split by fibrinolysis resistance grade, SOFA-2 tertile and ICU outcome.

|  | All | Fibrinolysis grade |  |  |  | SOFA-2 tertile |  |  |  | ICU outcome |  |  |
| --- | --- | --- | --- | --- | --- | --- | --- | --- | --- | --- | --- | --- |
| Protein<br>(units)<br>{normal<br>values<sup>c</sup>} | (283) | Grade 1<br>(118) | Grade 2<br>(71) | Grade 3<br>(94) | p<sup>a</sup> | ≤10<br>(107) | 11–13<br>(83) | ≥14<br>(93) | p<sup>a</sup> | Survived<br>(176) | Died<br>(107) | p<sup>b</sup> |
| tPA activity<br>(IU/mL)<br>{0.27 IU/mL} | 0.19<br>[0.07–<br>0.35] | 0.20<br>[0.08–<br>0.29] | 0.16<br>[0.05–<br>0.71] | 0.18<br>[0.07–<br>0.36] | 0.37 | 0.16<br>[0.07–<br>0.36] | 0.26<br>[0.05–<br>0.34] | 0.20<br>[0.07–<br>0.37] | 0.59 | 0.20<br>[0.07–0.37] | 0.18<br>[0.06–0.33] | 0.52 |
| PAI-1 activity<br>(IU/mL)<br>{1–40 IU/mL} | 37<br>[11–114] | 14<br>[7.1–42] | 35<br>[12–91] | 131<br>[51–300] | <0.001 | 17<br>[8.1–47] | 27<br>[8.3–77] | 117<br>[45–300] | <0.001 | 24<br>[8.5–67] | 105<br>[19–145] | <0.001 |
| Plasminogen<br>(µg/mL)<br>{77–168 µg/L} | 98<br>[64–223] | 131<br>[80–334] | 104<br>[64–229] | 77<br>[48–156] | <0.001 | 108<br>[75–289] | 109<br>[67–230] | 79<br>[50–215] | <0.001 | 103<br>[66–237] | 96<br>[62–212] | 0.41 |
| α2AP<br>(µg/mL)<br>{70 µg/L} | 59<br>[43–75] | 59<br>[47–77] | 63<br>[50–78] | 52<br>[36–70] | 0.22 | 63<br>[49–86] | 58<br>[45–75] | 53<br>[35–68] | 0.35 | 61<br>[45–77] | 56<br>[39–71] | 0.11 |

Data are median [interquartile range] of 283 samples in 69 patients with a median of 4 sampling episodes per patient. Fibrinolysis-resistance grade and SOFA-2 tertile are concurrent to the plasma sampling episode.
<sup>a</sup> p-values are linear-trend tests across ordered categories (grade 1→3; SOFA-2 tertile 1→3) from linear mixed-effects models on natural-log-transformed protein with a random patient intercept.
<sup>b</sup> p-values are two-sided Mann-Whitney U test.
tPA, tissue plasminogen activator; PAI-1, plasminogen-activator inhibitor-1; α2AP, α2-antiplasmin; SOFA, Sequential Organ Failure Assessment
<sup>c</sup> normal values obtained or derived from literature included with specific ELISA kits.

**Table 3.** Associations of plasma fibrinolysis proteins with fibrinolysis resistance and organ-failure severity.

| Protein | Fibrinolysis resistance (per e-fold increase in TPA-LT/FIBA10) |  |  | Organ failure (per SOFA-2 point) |  |
| --- | --- | --- | --- | --- | --- |
| | $\beta$ (95% CI) | Fold-change (95% CI) | p | Fold-change (95% CI) | p |
| tPA activity (IU/mL) | 0.014 (-0.070 to 0.098) | 1.01 (0.93 to 1.10) | 0.74 | 1.000 (0.976 to 1.024) | 0.97 |
| PAI-1 (IU/mL) | 0.570 (0.405 to 0.736) | 1.77 (1.50 to 2.09) | <0.001 | 1.228 (1.174 to 1.285) | <0.001 |
| PAI-1/tPA ratio | 0.426 (0.237 to 0.614) | 1.53 (1.27 to 1.85) | <0.001 | 1.241 (1.183 to 1.302) | <0.001 |
| Plasminogen (μg/mL) | -0.205 (-0.269 to -0.140) | 0.81 (0.76 to 0.87) | <0.001 | 0.946 (0.928 to 0.963) | <0.001 |
| α2AP (μg/mL) | 0.022 (-0.031 to 0.074) | 1.02 (0.97 to 1.08) | 0.42 | 0.992 (0.977 to 1.007) | 0.28 |
| α2AP/plasminogen ratio | 0.242 (0.171 to 0.313) | 1.27 (1.19 to 1.37) | <0.001 | 1.051 (1.029 to 1.073) | <0.001 |

Data are the linear mixed-effects model of the natural-log-transformed protein concentration on the stated predictor, with a random intercept for patient to account for repeated measurements (283 observations from 69 patients). $\beta$ is the change in log-protein per unit increase in the predictor. Fold-change = $\exp(\beta)$ , i.e. the multiplicative change in protein concentration per e-fold (~2.72-fold) increase in TPA-LT/FIBA10, or per one-point increase in SOFA-2. A fold-change above 1 indicates a rising concentration; below 1, a falling concentration. The 95% confidence intervals are stated within brackets. TPA-LT/FIBA10, tPA-enhanced viscoelastometric lysis time normalised to the fibrin-specific clot amplitude (higher values indicate greater fibrinolysis resistance); SOFA, Sequential Organ Failure Assessment; tPA, tissue plasminogen activator; PAI-1, plasminogen-activator inhibitor-1; α2AP, α2-antiplasmin.

### Principal component analysis and unsupervised clustering

When the four fibrinolysis proteins were analysed jointly with SOFA-2 and log(TPA-LT/FIBA10), the dominant axis of variation (PC1) accounted for 35.3% of the total variance (Figure 2A, 269 observations in 67 patients). This is more than the 16.7% expected by chance with six variables, indicating substantial shared structure. PAI-1 (+0.50), log(TPA-LT/FIBA10) (+0.45) and SOFA-2 (+0.43) loaded positively on this axis, while plasminogen (−0.46) and α2AP (−0.40) loaded negatively. A negligible contribution was made by tPA activity (+0.04). Observations from Grade 3 patients and worse organ failure distributed toward the positive end of this axis, and Grade 1 observations and less organ failure toward the negative end. The second axis (PC2, 21.6% of variance) in opposition and independent of PC1, shown by its orthogonal orientation, carried positive loadings for the fibrinolysis proteins dominated by α2AP (+0.61) and plasminogen (+0.51). Plasminogen and α2AP loaded together on PC2 because their concentrations were strongly correlated (ρ=+0.62, p<0.001) (Figure 2B), while α2AP was not independently associated with fibrinolysis resistance (ρ=−0.07, p=0.23, Figure 2B). This therefore reflected overall plasma protein abundance but no fibrinolytic contrast. No other significant correlations were found between plasminogen and albumin, plasmin-antiplasmin complex or D-dimer, while a weak inverse relationship was found with PAI-1 (Figure S3, Additional file 1). Unsupervised Ward clustering, performed without reference to ICU outcome, grouped the 269 observations into an inhibitor-high, substrate-depleted cluster (79 observations; higher PAI-1, lower plasminogen) and a preserved-fibrinolysis cluster (190 observations, higher plasminogen), separating principally along PC1 (Figure 2C). The ICU mortality was 61% among patients whose observations fell predominantly in the inhibitor-high, substrate depleted cluster *versus* 29% in the preserved-fibrinolysis cluster (p=0.022). The fibrinolysis resistance axis represented by PC1 increased with grade (ρ=+0.62, p<0.001) and was increased in ICU non-survivors compared to survivors (p<0.001) (Figure 2D). Taken together, plasma fibrinolysis protein derangement dominated by increased PAI-1 and reduced plasminogen, organ-failure severity and fibrinolysis resistance grade were thus coupled aspects of the common septic insult.

**Figure 2.**
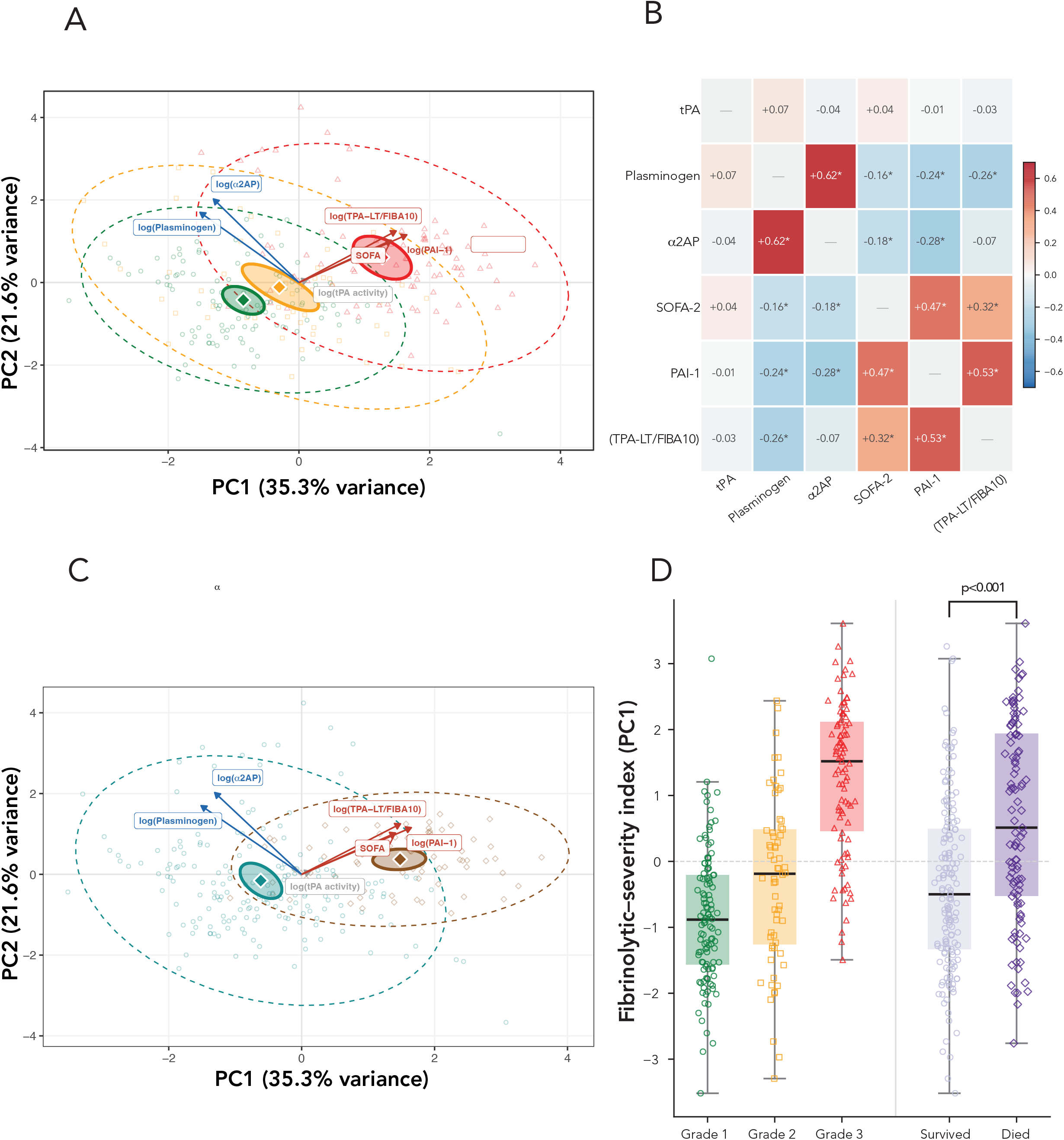
Integrated structure of plasma fibrinolysis proteins, organ-failure severity and fibrinolysis resistance measured bedside by tPA-enhanced VET. Principal component analysis (PCA) was performed on six standardised variables: log(tPA activity), log(PAI-1), log(plasminogen), log(α2AP), SOFA-2 and log(TPA-LT/FIBA10) to capture their joint variation. (**A**) PC1–PC2 biplot with observations coloured by fibrinolysis resistance grade (Grade 1 green, Grade 2 amber, Grade 3 red). Arrows are variable loadings, dashed ellipses are 95% data (concentration) ellipses and solid filled ellipses are 95% confidence regions for each group centroid. Arrows pointing the same way represent variables that change together. All of log(TPA-LT/FIBA10), log(PAI-1) and SOFA-2 loaded positively on PC1. The minimally short grey arrow for tPA indicates its poor joint variability. PC2 is orthogonal to PC1, ie. statistically independent, with log(plasminogen) loading together with log(α2AP). Note that loading in the PCA does not constitute an association (shown in panel B). The joint loading for α2AP and plasminogen is explained by their correlation (ρ = +0.62, panel B), while α2AP had no independent relationship with resistance (ρ = −0.07, not significant, panel B). (**B**) Pairwise Spearman correlation matrix for the same six variables as in the PCA (asterisks denote p < 0.05. (**C**) The PCA re-coloured by unsupervised Ward k = 2 cluster assignment, which was derived from the six variables alone without reference to grade or outcome. (**D**) The PC1 score, interpretable as a composite fibrinolytic–severity index, plotted by fibrinolysis grade and by ICU outcome.

### Temporal trajectories of fibrinolysis proteins

To capture the clinical impact of fibrinolysis proteins over time, their trajectories were assessed by the interaction of sampling episode and SOFA-2 tertile and ICU survival (Figure 3). Statistical inference was restricted to episodes 1–3, where group sizes were adequate. Only PAI-1 and plasminogen are shown since tPA and α2AP showed no correlation with organ failure severity (see Figure S2A, Additional file 1). The trajectories for PAI-1 levels in both the highest SOFA-2 tertile and non-survivors showed no early change (episode×SOFA-2 interaction p=0.32; episode×ICU outcome interaction p=0.87). In contrast, PAI-1 levels in the low and middle SOFA-2 tertiles as well as in survivors showed an early, progressive decrease (episode×SOFA-2 interaction p=0.03; episode×ICU outcome interaction p<0.001). No significant differences in the early trajectories were found for plasminogen (episode×SOFA-2 interaction p=0.89; episode×ICU outcome interaction, p=0.26). The late apparent increase in plasminogen levels for patients in the lowest SOFA-2 tertile and ICU survivors was not formally tested given the sparse data due to deaths and discharges, thus is descriptive only.

**Figure 3.**
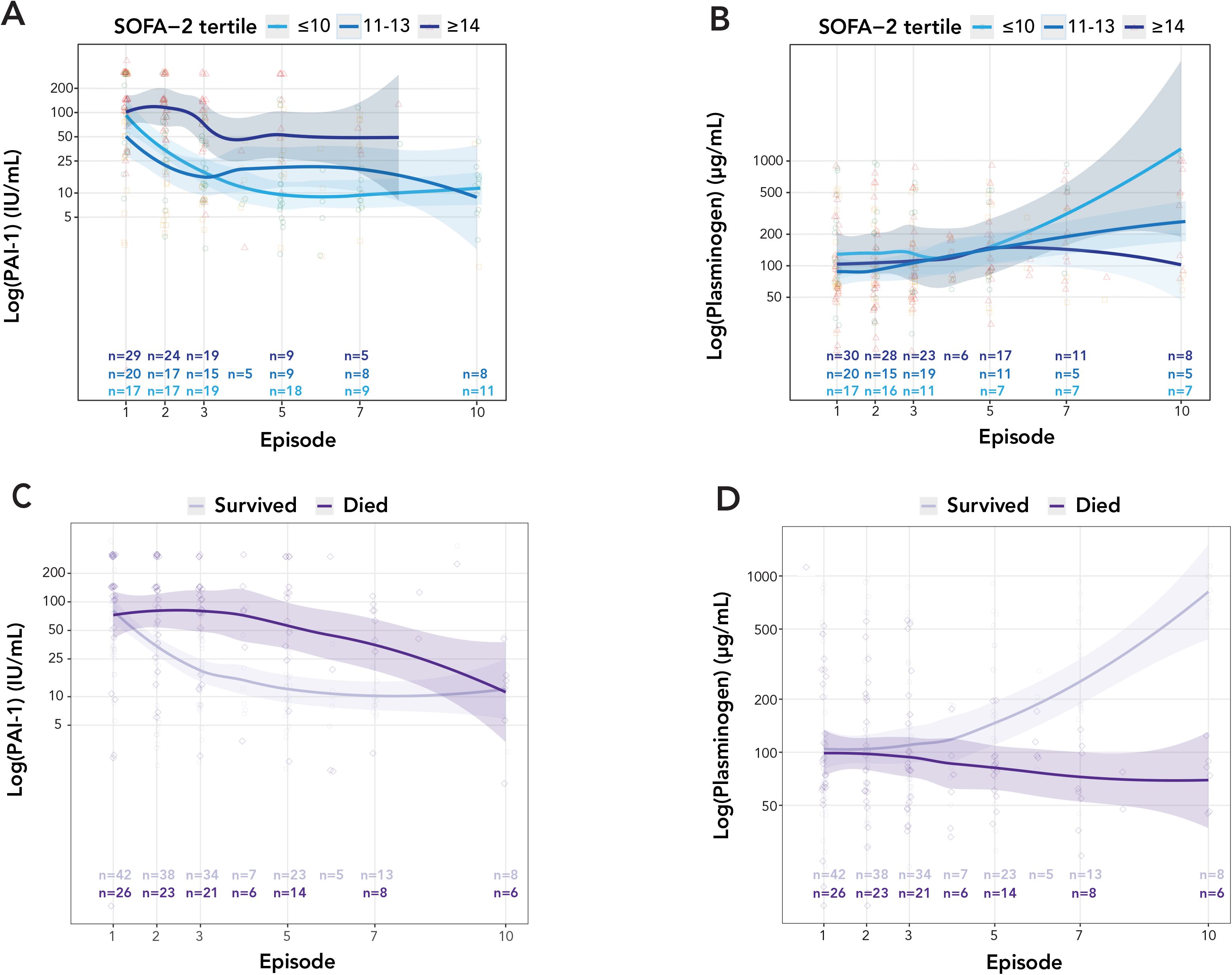
Serial PAI-1 and plasminogen trajectories by organ-failure severity and ICU outcome. PAI-1 (A, C) and plasminogen (B, D) are shown across sampling episodes, stratified by SOFA-2 tertile (A, B) and by ICU outcome (C, D). The trajectory lines show LOESS smoothing with the 95% confidence interval shaded. The number of contributing observations per group per episode is annotated on each panel. Sampling data became progressively sparser at later episodes, and trajectories beyond the early episodes are descriptive and hypothesis-generating, also considering their inherent survival bias.

### Sensitivity analyses

All conclusions based on observational level data remained unchanged using patient level data incorporating within and between patient variability for the correlations between fibrinolysis proteins and TPA-LT/FIBA10 and jointly in the PCA (see Figures S2-S4, Additional file 1). Furthermore, all associations retained their direction and statistical significance excluding the five patients (see Table 1) on therapeutic anticoagulation with unfractionated heparin (median 15000 [IQR 15000-22459] U/day) (see Table S2, Additional file 1).

## DISCUSSION

This observational study in a cohort of patients admitted to ICU with sepsis demonstrated (i) that the bedside VET measured fibrinolysis resistance parameter, TPA-LT/FIBA10, plasma fibrinolysis proteins, PAI-1 and plasminogen, and organ-failure severity are inter-related, varying in coordination rather than independently; (ii) a phenotype characterised by plasma fibrinolysis protein inhibitor-high, substrate-depleted, predominantly Grade 3 fibrinolysis resistance had an ICU mortality more than double that of patients with preserved fibrinolysis proteins and predominantly Grade 1 and 2 fibrinolysis resistance. (iii) During the ICU stay, PAI-1 remained persistently elevated throughout in patients with severe organ failure and in non-survivors, while decreasing over time in patients with less severe organ failure and in survivors. Plasminogen levels were low on admission and tended to increase only in patients with less organ failure and in survivors, while this observation had insufficient statistical power.

### Associations between fibrinolysis proteins, organ failure and VET fibrinolysis resistance

The spline and linear correlations between TPA-LT/FIBA10 and PAI-1 and plasminogen coincided within their confidence bands throughout, indicating continuous, monotone protein–fibrinolysis resistance relationships without any thresholds at the fibrinolysis resistance grade transitions. This suggests a common pathophysiological axis of worsening driven by inhibition of fibrinolysis pathways. The grading system of fibrinolysis resistance based on TPA-LT/FIBA10 [4] has been designed to facilitate clinical implementation and interpretability of this specific feature of sepsis induced coagulopathy. It enables pragmatic stratification for study enrichment or interventions. The concordance between fibrinolysis protein concentrations and both the VET-derived resistance grade and tertiles of organ-failure severity provides biological support for this bedside test. The same high PAI-1, low plasminogen pattern that characterised greater SOFA-2 scores was also reflected in the fibrinolysis resistance grades. This indicates that the fibrinolysis resistance grade captures a clinically meaningful protein signature.

### Principal component analysis and unsupervised clustering

The findings in this study align with the accepted pathophysiology of sepsis-induced coagulopathy, in which impaired fibrinolysis correlates with organ failure and is driven by sustained increase in PAI-1 released from endothelial cells, platelets and inflammatory macrophages [2, 21, 22]. The dominance of PAI-1 on the PCA integrated axis, and its concurrent correlation with fibrinolysis resistance quantified by TPA-LT/FIBA10, place inhibitor excess at the centre of the resistant phenotype [9]. The parallel depletion of plasminogen, however, is equally compelling and consistent with the emerging concept of a quantitative substrate deficiency [1, 23]. The observed levels on ICU admission in Grade 3 resistant patients, around 100 µg/mL, are associated with fibrinolysis deficiency [13, 15]. The biologically activatable levels of plasminogen could potentially be even lower than the ELISA antigen levels reported here [24]. There are several documented causes of plasminogen deficiency in sepsis [25]. Depletion may occur during an early phase of hyperfibrinolysis [8], however, we have not observed this state in our ICU population using viscoelastic testing and found no correlation between plasminogen deficiency and plasmin:antiplasmin (PAP) complexes or D-dimers. It is likely, therefore, that any hyperfibrinolytic state occurs prior to the onset of organ dysfunction, the principal trigger for ICU admission. Compromised hepatic function may result in reduced plasminogen production [25], which is a plausible contributor in our cohort given the associations between reduced plasminogen levels and increased organ failure SOFA-2 score. Immune cell migration and tissue repair and remodelling processes increase plasminogen consumption and are highly likely to be occurring in our severe sepsis population [26, 27]. Additionally, neutrophil extracellular trap production results in plasminogen cleavage by neutrophil elastase into fragments, and the release of DNA, both of which competitively inhibit fibrin binding by intact plasminogen [15, 28–31]. All these processes potentially contribute to a reduction in the functional substrate available for plasmin generation despite ongoing fibrin formation [1, 31]. A proteomic analysis of sepsis patient plasma could reveal specific proteins and biological pathways that associate with plasminogen reductions and Grade 3 fibrinolysis resistance.

That tPA activity showed no gradient is coherent with the recognised biphasic response, in which an early transient rise in tPA is rapidly eclipsed by sustained PAI-1 elevation [1, 32, 33]. By the time of the first blood sample in ICU within 24 hours of admission, the fibrinolytic state was determined by increased inhibitor and low substrate, rather than by the activity of tPA per se, that did not discriminate severity. The convergence of anti-fibrinolytic changes in fibrinolysis proteins with SOFA-2 onto one axis supports the view that fibrinolytic insufficiency is an integral component of the organ-failure process associated with sepsis rather than an epiphenomenon. Furthermore, the concordance between an unguided fibrinolysis protein-based cluster analysis and the ICU outcomes substantiates that the plasma protein signatures reflect a clinically relevant process.

The PCA was performed on the log-transformed protein concentrations to capture their joint variation. Although plasminogen and α2-antiplasmin loaded in the same direction on PC1 this alignment reflects their strong mutual correlation (ρ=+0.62) rather than equivalent biological roles. This is not surprising given that both proteins are synthesised in the liver.

### Fibrinolysis protein trajectories

The changes in PAI-1 and plasminogen over time carry their own implications for fibrinolysis resistance in sepsis. Elevated PAI-1 levels were already established at admission to ICU. Importantly, the levels remained unchanged over the first 3 episodes (72 hours) in patients with the worst organ failure scores and non-survivors but decreased progressively in patients with less severe organ failure and who survived ICU. The dynamic biological signal relating to fibrinolysis resistance was thus present early in the clinical course of sepsis, with failure to reduce PAI-1 associated with worse outcomes, similar to a previous report of progressively increasing PAI-1 in non-survivors [34]. It raises the possibility that dynamic, not merely baseline, inhibitor burden carries prognostic information and that serial monitoring could refine risk stratification. In the context of any fibrinolysis interventions, this favours early bedside phenotyping and an early potential therapeutic window before sustained fibrin deposition in vital organs, progressive plasminogen depletion, and end-organ injury have advanced beyond a reversible threshold. Plasminogen levels were low across all SOFA-2 tertiles and in both ICU survivors and non-survivors on admission to ICU. Interestingly, plasminogen levels tended to increase beyond five days of ICU admission in patients in the lowest SOFA-2 tertile and ICU survivors. While the trajectory model is acknowledged to be underpowered for formal inference beyond the first three episodes, it is nonetheless biologically plausible.

Persistently elevated PAI-1 and low plasminogen levels in patients in whom the septic process fails to resolve impose a progressively worsening constraint on fibrinolysis capacity. This corresponds to the observation that patients with the worst (Grade 3) fibrinolysis resistance have a greater than 70% probability to remain in that state, which is associated with a rapid increase in mortality over the first 5-7 days of ICU admission as previously reported [4]. The early evolution of PAI-1 and plasminogen levels are aligned with the recent discussion of traditional randomised controlled trials in sepsis failing since patient heterogeneity and dynamic trajectories were not recognised [35]. Study enrichment by fibrinolytic phenotype and by timing, using a rapid bedside VET grading assay substantiated by the underlying protein biology, may provide a more tractable approach compared to enrolment based on sepsis diagnosis alone.

### Future directions

The plasma fibrinolysis protein data suggest a biological rationale for targeted potential profibrinolytic strategies in sepsis. Any such interventions need to be phenotype-directed rather than uniform, for example supplemental plasminogen as a substrate-oriented approach would pragmatically be reserved for Grade 3 fibrinolysis resistance where the most effect can be expected. The dominance of PAI-1 inhibition, with a uniformly low tPA activity, represents an overwhelmed activator pathway. This, together with diminishing plasminogen levels, as correlates of fibrinolysis resistance, provide a mechanistic argument that the therapeutic target in severe resistance is not only the activator arm of the fibrinolytic system but also the constraint on functional substrate downstream. The correlation between plasma fibrinolysis proteins and the fibrinolysis resistance grades suggest that the optimal intervention point may be identifiable by performing bedside tPA-enhanced VET. A recent pilot trial of plasminogen-containing plasma supplementation in sepsis-induced coagulopathy, which significantly raised functional plasminogen and plasmin generation without altering PAI-1 or tPA [23], directly addresses the substrate depletion signal that this study characterised at the protein level. The directional concordance between the pilot trial and the protein signatures reported here is mechanistically coherent. Notably, the pilot trial identified the absence of kinetic fibrinolytic sampling as a key limitation. The present study using serial measurements begin to fill that gap by characterising how levels of relevant substrate and inhibitor proteins behave over the early course of ICU admission, including correlations to the bedside VET quantification of fibrinolysis resistance by the TPA-LT/FIBA10 variable.

### Strengths and limitations

This study has several limitations. It is a single-centre observational study with a pilot sample size. The correlation, clustering and PCA analyses treat repeated within-patient observations as independent and are therefore presented as exploratory descriptions of the study cohort. While the associations between fibrinolysis proteins, VET-derived fibrinolysis resistance and organ failure severity as the primary outcome were congruent on both observational and patient levels, the ICU survival analyses were secondary and should be regarded as hypothesis-generating. As an observational study, the associations reported cannot be used to infer any causality. Other unmeasured modulators of fibrinolysis, such as TAFI, may also contribute to the association with organ failure. Additionally, the plasminogen ELISA assay may have overestimated the functional plasminogen pool with a functional assay required to resolve this issue in future studies. Finally, formal trajectory statistical inference was deliberately restricted to the first three sampling episodes where observations generated adequate statistical power, whereas later time points should be viewed as descriptive.

In conclusion, plasma fibrinolysis protein derangement, organ-failure severity and bedside fibrinolysis resistance in sepsis are facets of a single, graded fibrinolytic-insufficiency phenotype that is established early and dominated by PAI-1 excess with reduced plasminogen levels. This integrated view provides a biochemical foundation for the bedside VET-derived quantification of fibrinolysis resistance and its grading, suggests targets for a potential substrate-dependent reversibility of severe resistance, and supports the prospective, timing-sensitive evaluation of phenotype-directed profibrinolytic strategies.

## Data Availability

The datasets generated and analysed during the current study are not publicly available due to restrictions by the Human Research Ethics Committee but are available from the corresponding author on reasonable request and following ethics approval.

## LIST OF ABBREVIATIONS

APACHE III: Acute Physiology and Chronic Health Evaluation III
α2AP: α2-antiplasmin
DIC: Disseminated intravascular coagulation
ELISA: Enzyme-linked immunosorbent assay
FIBA10: Fibrin-specific clot amplitude at 10 minutes
LOESS: Locally estimated scatterplot smoothing
NETs: Neutrophil extracellular traps
PAI-1: Plasminogen activator inhibitor-1
PC1/PC2: First / second principal component
PCA: Principal component analysis
SIC: Sepsis-induced coagulopathy
SOFA-2: Sequential Organ Failure Assessment (version 2)
tPA: tissue plasminogen activator
TPA-LT: Tissue plasminogen activator-challenged lysis time
TPA-LT/FIBA10: Fibrin-adjusted clot lysis time (TPA-LT normalised to FIBA10)
VET: Viscoelastometric testing

## DECLARATIONS

## Ethics approval and consent to participate

This is a substudy of the FORTITUDE single-centre observational study in sepsis (NCT06680180) conducted at Liverpool ICU, Australia, and approved by the South Western Sydney Local Health District Human Research Ethics Committee (2022/ETH02122) with a waiver of consent as permitted under the National Statement on Ethical Conduct in Human Research 2023 for low-risk research in critically ill patients.

## Consent for publication

Not applicable

## Authors’ contributions

LC: Project conception, design, funding, conduct, data collection, analyses and manuscript preparation. MS: Patient screening, data collection and manuscript preparation. ES: Patients, screening, data collection and database management. RL: Intellectual input and manuscript preparation. PC: Intellectual input and manuscript preparation. RM: Data analyses and ELISA assay supervision, intellectual input and manuscript preparation. CK: Data analyses and ELISA assay supervision, intellectual input and manuscript preparation. SF: Data analyses and manuscript preparation. PKM: Intellectual input and manuscript preparation. AA: Project conception, design, conduct, clinical supervision, data collection, analyses and manuscript preparation.

## Acknowledgements

The authors extend their sincere gratitude to the patients and their families who participated in this study. We are grateful to the nursing and medical staff of the Liverpool Hospital Intensive Care Unit for their assistance with this study, in particular the ClotPro Champions, Intensive Care nurses who were trained on the ClotPro technology and provided bedside capability: Alexandra Heath, Emma Hill, Marcus Iedani, Karla Lopez, Elisa McClure, Jennifer McClure, Yvette McManus, Carlos Mendoza, Jennene Miller, Mostafa Hasha, Sharyn Parker, Julio Ruiz Davila, Krystle-Lee Strickland, Anne Tran, Bija Unnikrishnan, Harley Whitfield, Sanjay Singh, Katie Mills. Joanne Chia, Tammy Lam, Zikou Liu for performing the ELISA assays. Mrs Korina Valentine, consumer representative, provided essential input to the project design as a survivor of sepsis.

## Additional file 1

**Table S1.**
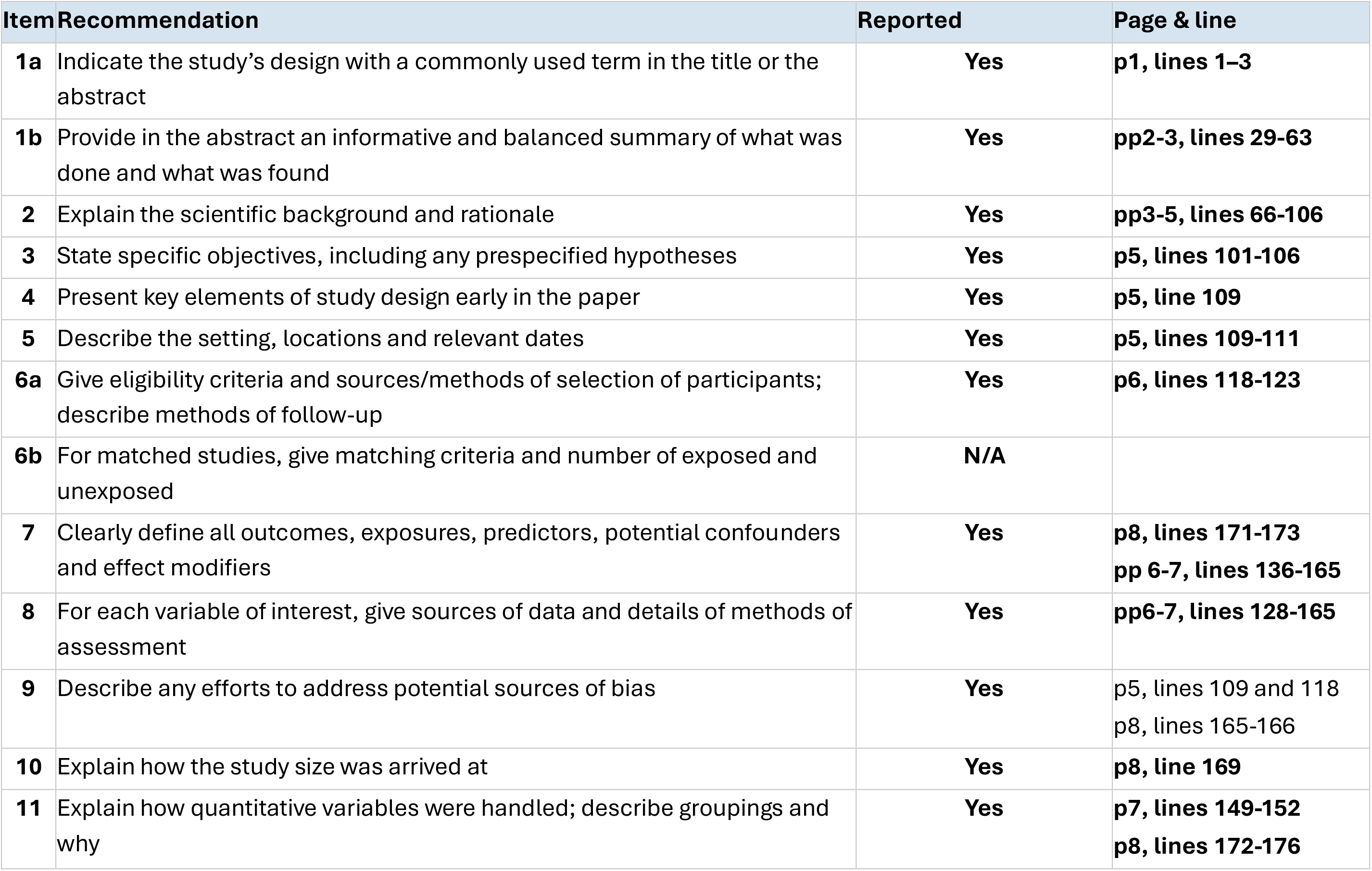
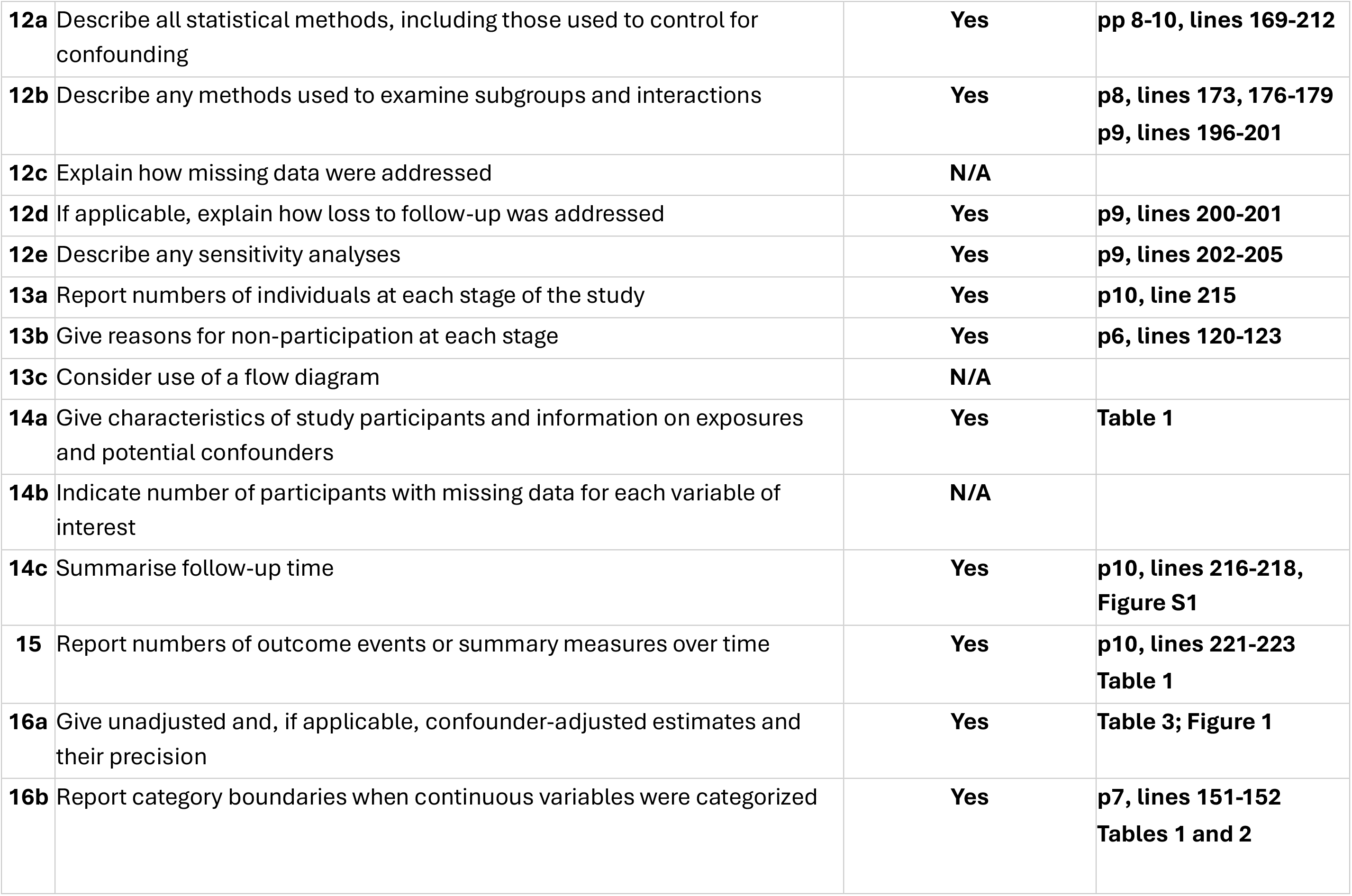
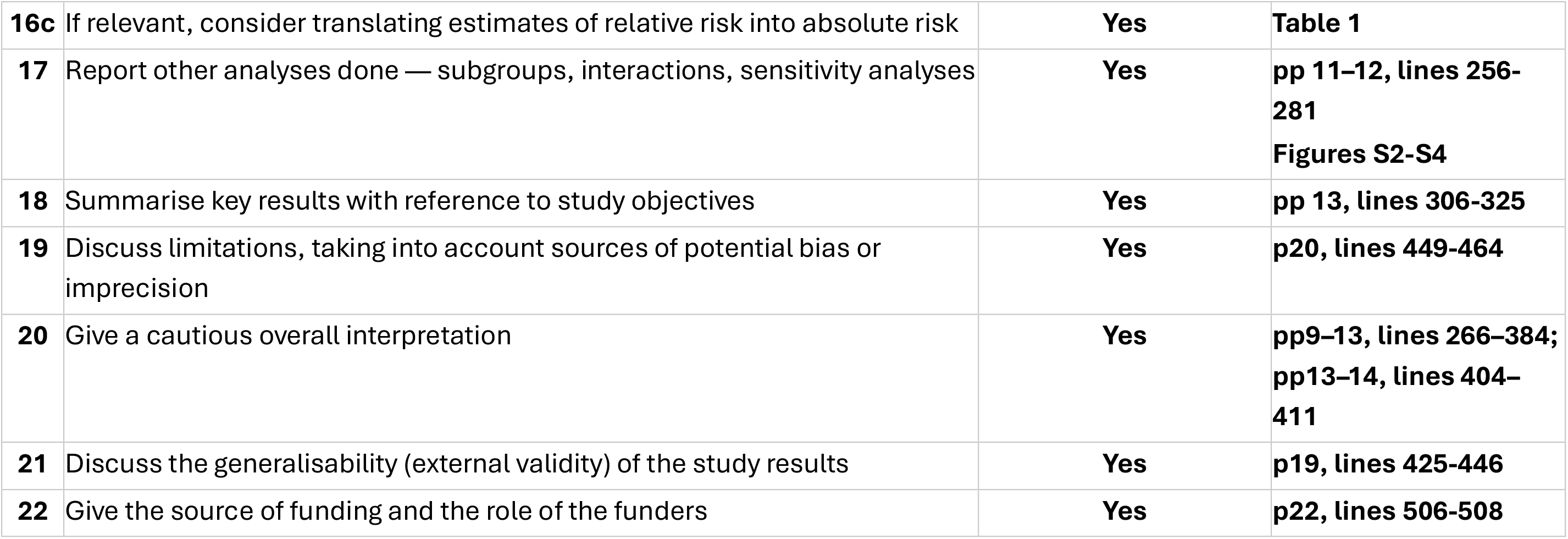
STROBE checklist. Page and line numbers refer to the submitted Word document containing the text body of the manuscript.

| **Item** | **Recommendation** | **Reported** | **Page & line** |
| --- | --- | --- | --- |
| **1a** | Indicate the study's design with a commonly used term in the title or the abstract | **Yes** | **p1, lines 1–3** |
| **1b** | Provide in the abstract an informative and balanced summary of what was done and what was found | **Yes** | **pp2-3, lines 29-63** |
| **2** | Explain the scientific background and rationale | **Yes** | **pp3-5, lines 66-106** |
| **3** | State specific objectives, including any prespecified hypotheses | **Yes** | **p5, lines 101-106** |
| **4** | Present key elements of study design early in the paper | **Yes** | **p5, line 109** |
| **5** | Describe the setting, locations and relevant dates | **Yes** | **p5, lines 109-111** |
| **6a** | Give eligibility criteria and sources/methods of selection of participants; describe methods of follow-up | **Yes** | **p6, lines 118-123** |
| **6b** | For matched studies, give matching criteria and number of exposed and unexposed | **N/A** |  |
| **7** | Clearly define all outcomes, exposures, predictors, potential confounders and effect modifiers | **Yes** | **p8, lines 171-173**<br>**pp 6-7, lines 136-165** |
| **8** | For each variable of interest, give sources of data and details of methods of assessment | **Yes** | **pp6-7, lines 128-165** |
| **9** | Describe any efforts to address potential sources of bias | **Yes** | p5, lines 109 and 118<br>p8, lines 165-166 |
| **10** | Explain how the study size was arrived at | **Yes** | **p8, line 169** |
| **11** | Explain how quantitative variables were handled; describe groupings and why | **Yes** | **p7, lines 149-152**<br>**p8, lines 172-176** |
| **12a** | Describe all statistical methods, including those used to control for confounding | **Yes** | **pp 8-10, lines 169-212** |
| **12b** | Describe any methods used to examine subgroups and interactions | **Yes** | **p8, lines 173, 176-179**<br>**p9, lines 196-201** |
| **12c** | Explain how missing data were addressed | **N/A** |  |
| **12d** | If applicable, explain how loss to follow-up was addressed | **Yes** | **p9, lines 200-201** |
| **12e** | Describe any sensitivity analyses | **Yes** | **p9, lines 202-205** |
| **13a** | Report numbers of individuals at each stage of the study | **Yes** | **p10, line 215** |
| **13b** | Give reasons for non-participation at each stage | **Yes** | **p6, lines 120-123** |
| **13c** | Consider use of a flow diagram | **N/A** |  |
| **14a** | Give characteristics of study participants and information on exposures and potential confounders | **Yes** | **Table 1** |
| **14b** | Indicate number of participants with missing data for each variable of interest | **N/A** |  |
| **14c** | Summarise follow-up time | **Yes** | **p10, lines 216-218,**<br>**Figure S1** |
| **15** | Report numbers of outcome events or summary measures over time | **Yes** | **p10, lines 221-223**<br>**Table 1** |
| **16a** | Give unadjusted and, if applicable, confounder-adjusted estimates and their precision | **Yes** | **Table 3; Figure 1** |
| **16b** | Report category boundaries when continuous variables were categorized | **Yes** | **p7, lines 151-152**<br>**Tables 1 and 2** |
| **16c** | If relevant, consider translating estimates of relative risk into absolute risk | **Yes** | **Table 1** |
| **17** | Report other analyses done — subgroups, interactions, sensitivity analyses | **Yes** | **pp 11–12, lines 256–281**<br>**Figures S2–S4** |
| **18** | Summarise key results with reference to study objectives | **Yes** | **pp 13, lines 306–325** |
| **19** | Discuss limitations, taking into account sources of potential bias or imprecision | **Yes** | **p20, lines 449–464** |
| **20** | Give a cautious overall interpretation | **Yes** | **pp9–13, lines 266–384;**<br>**pp13–14, lines 404–411** |
| **21** | Discuss the generalisability (external validity) of the study results | **Yes** | **p19, lines 425–446** |
| **22** | Give the source of funding and the role of the funders | **Yes** | **p22, lines 506–508** |

**Table S2.** Sensitivity analyses excluding patients on therapeutic anticoagulation. Five patients received therapeutic anticoagulation using unfractionated heparin, median dose 15000 [interquartile range 15000-22459] U/day, that are compared (Excluding) vs. the full cohort (All).

| Plasma proteins vs. log(TPA-LT/FIBA10) |  |  |  |  |
| --- | --- | --- | --- | --- |
| Protein | Spearman ρ (p): All | Excluding |  |  |
| tPA activity | -0.02 (0.784) | -0.00 (0.960) |  |  |
| PAI-1 | +0.54 (<0.001) | +0.51 (<0.001) |  |  |
| Plasminogen | -0.26 (<0.001) | -0.31 (<0.001) |  |  |
| α2AP | -0.08 (0.172) | -0.03 (0.578) |  |  |
| Linear-trend p-values across SOFA-2 tertile and fibrinolysis grade |  |  |  |  |
| Protein | SOFA-2 tertile: All | Excluding | Grade: All | Excluding |
| tPA activity | 0.588 | 0.587 | 0.366 | 0.372 |
| PAI-1 | <0.001 | <0.001 | <0.001 | <0.001 |
| Plasminogen | <0.001 | <0.001 | <0.001 | <0.001 |
| α2AP | 0.355 | 0.413 | 0.224 | 0.179 |

| Mixed-effects regression per e-fold change (95% CI) |  |  |  |  |
| --- | --- | --- | --- | --- |
| Protein | TPA-LT/FIBA10: All | Excluding | SOFA-2 point: All | Excluding |
| tPA activity | 1.02 (0.94–1.10) | 1.02 (0.94–1.10) | 1.00 (0.98–1.02) | 1.00 (0.98–1.02) |
| PAI-1 | 1.79 (1.51–2.11) | 1.74 (1.47–2.06) | 1.23 (1.17–1.28) | 1.23 (1.17–1.29) |
| Plasminogen | 0.82 (0.77–0.87) | 0.81 (0.76–0.86) | 0.95 (0.93–0.96) | 0.95 (0.93–0.97) |
| α2AP | 1.02 (0.97–1.07) | 1.03 (0.98–1.08) | 0.99 (0.98–1.01) | 0.99 (0.98–1.01) |
| Principal component analysis |  |  |  |  |
| Variable | PC1 All | PC1 Excluding | PC2 All | PC2 Excluding |
| tPA activity | +0.04 | +0.04 | +0.02 | +0.02 |
| PAI-1 | +0.50 | +0.49 | +0.35 | +0.36 |
| Plasminogen | -0.46 | -0.50 | +0.51 | +0.46 |
| α2AP | -0.40 | -0.39 | +0.61 | +0.63 |
| SOFA-2 | +0.43 | +0.41 | +0.31 | +0.33 |
| TPA-LT/FIBA10 | +0.45 | +0.43 | +0.38 | +0.39 |
| Clustering analysis |  |  |  |  |

| Analysis | Inhibitor-high | Preserved-fibrinolysis | p-value |
| --- | --- | --- | --- |
| All | 11/18 (61%) | 14/49 (29%) | 0.022 |
| Excluding | 9/15 (60%) | 14/49 (29%) | 0.035 |

**Figure S1.**
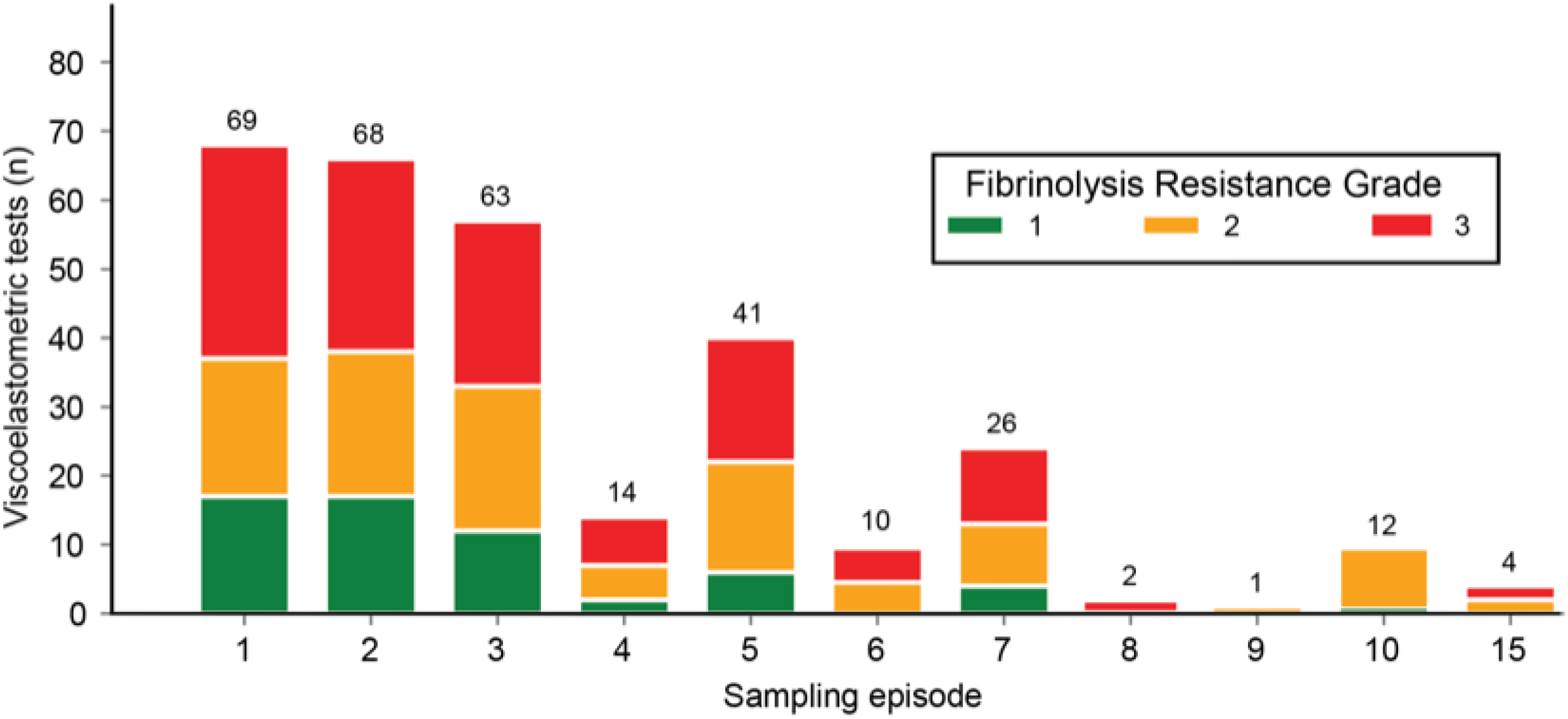
Data collections timepoints.

**Figure S2A.**
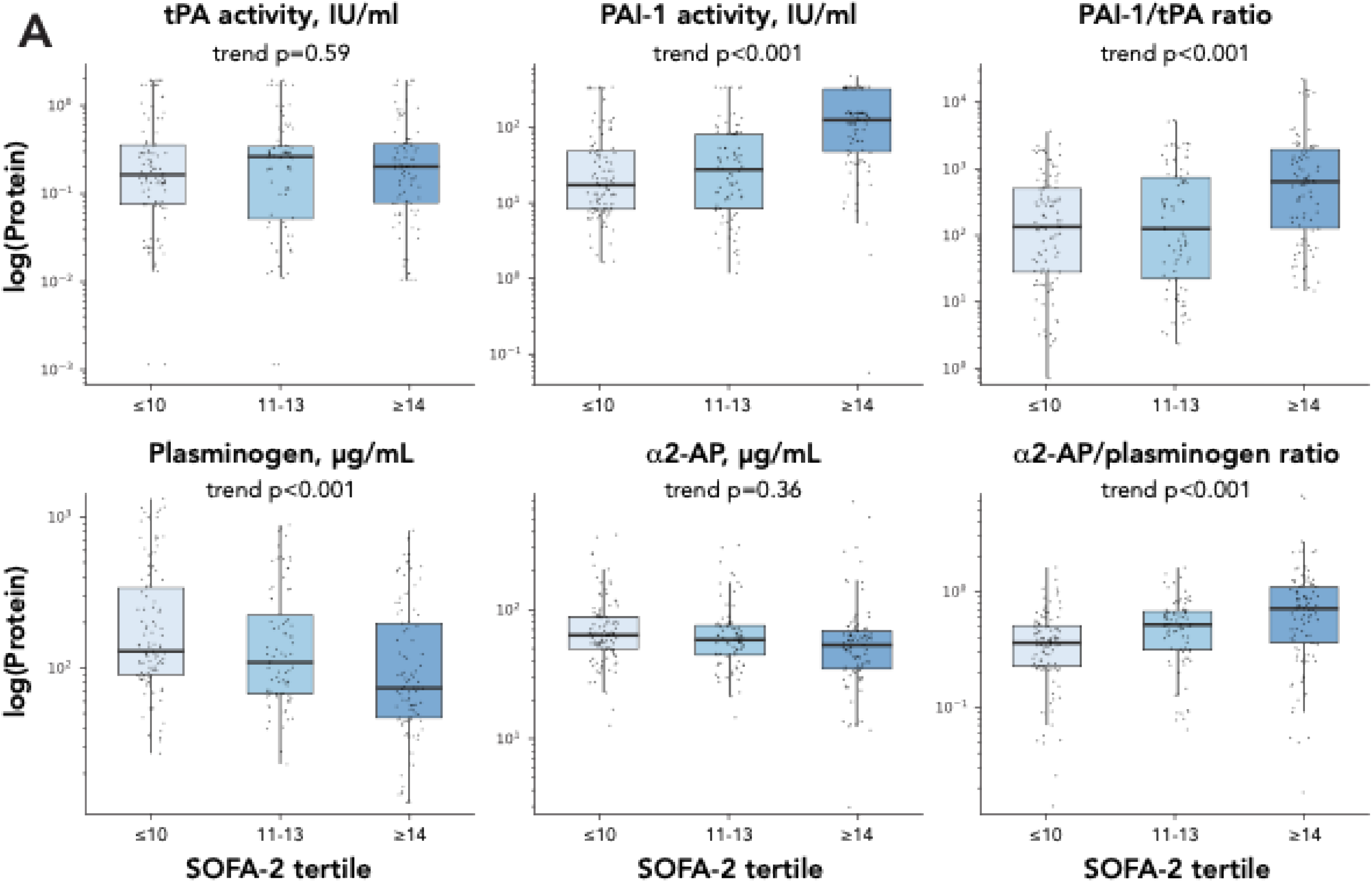
Correlations between fibrinolysis proteins and SOFA-2 score tertiles.

**Figure S2B.**
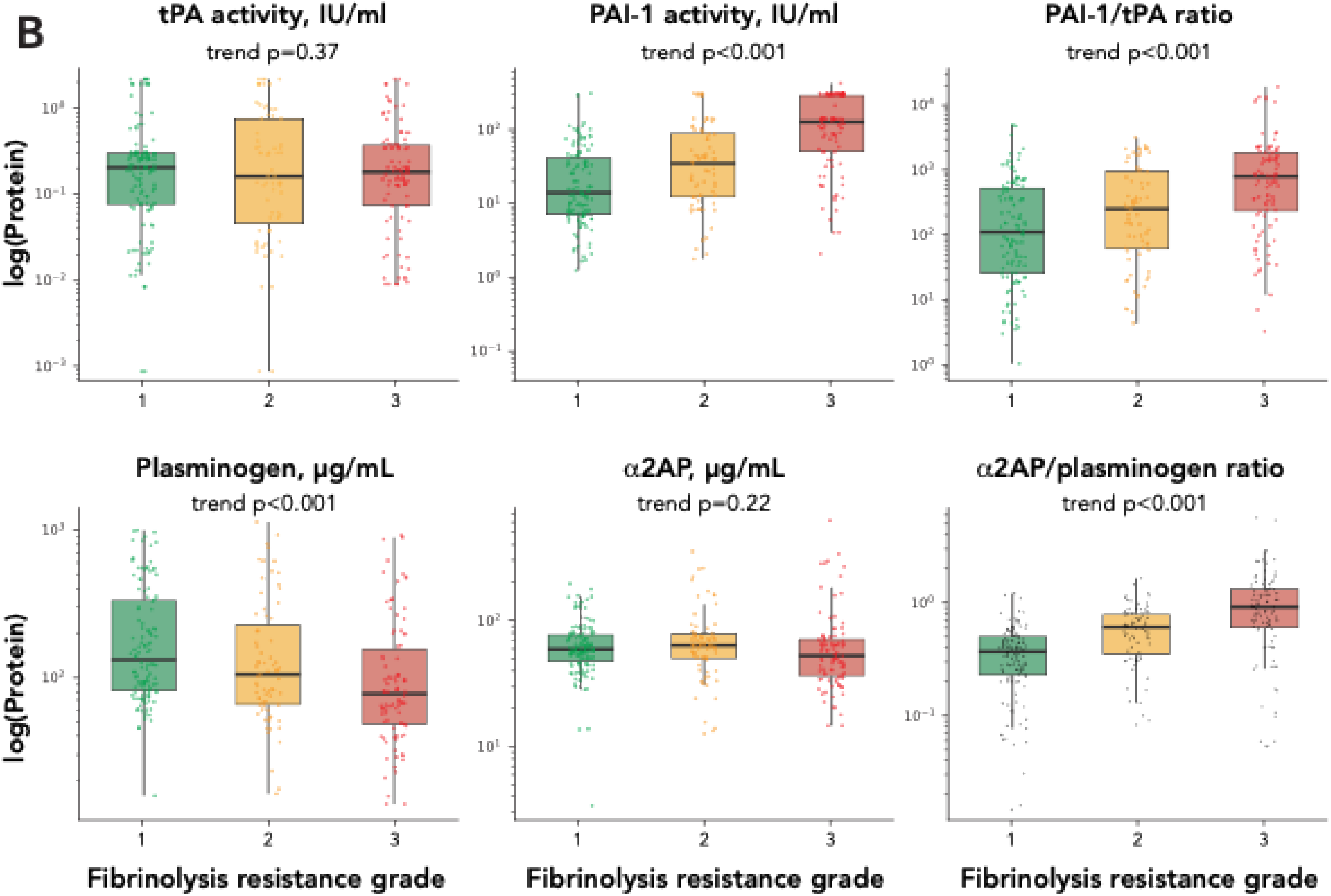
Correlations between fibrinolysis proteins and fibrinolysis resistance grades.

**Figure S3.**
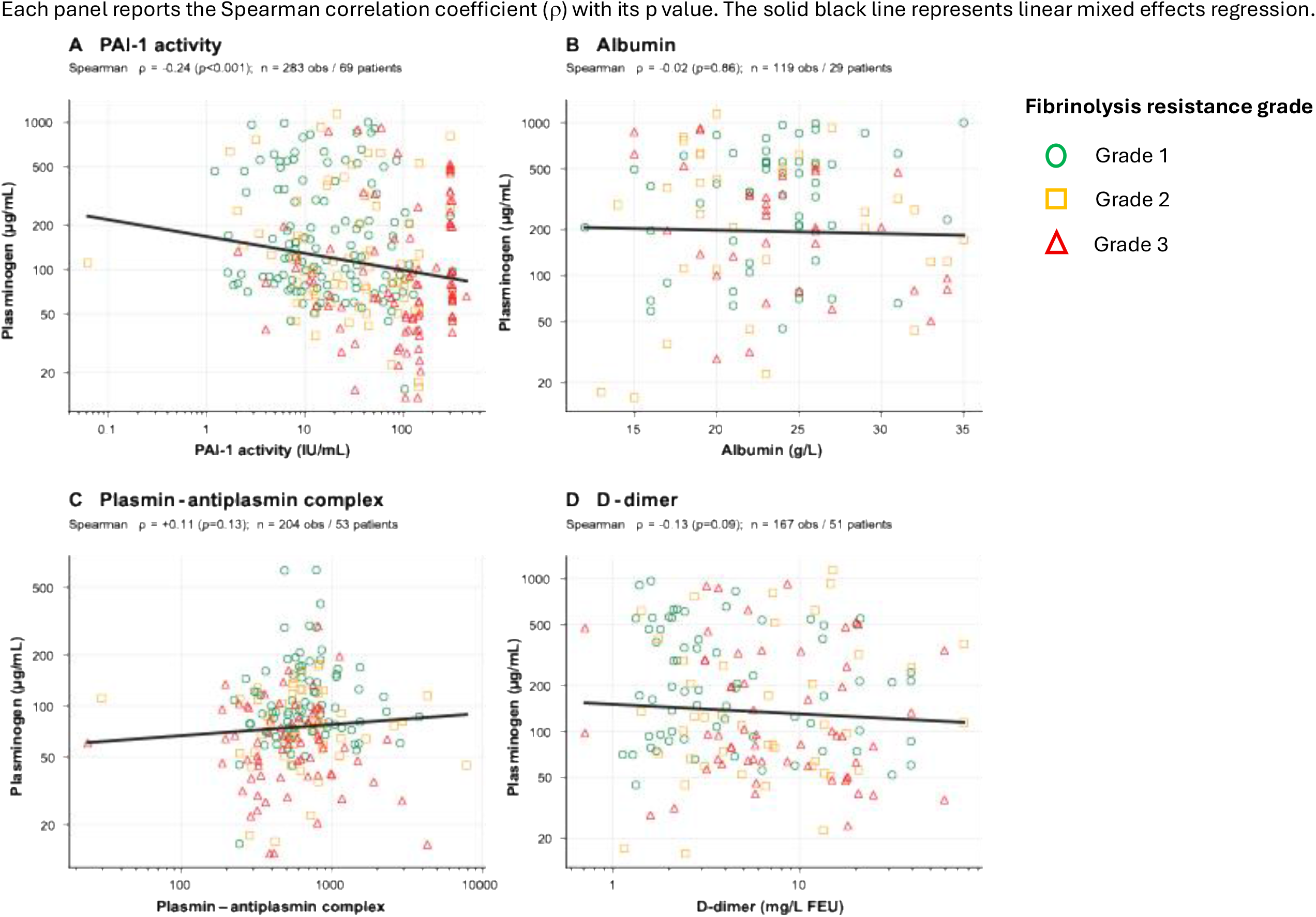
Correlations between plasminogen levels and albumin, PAP complex and D-dimer levels. Each panel reports the Spearman correlation coefficient (ρ) with its p value. The solid black line represents linear mixed effects regression.

**Figure S4.**
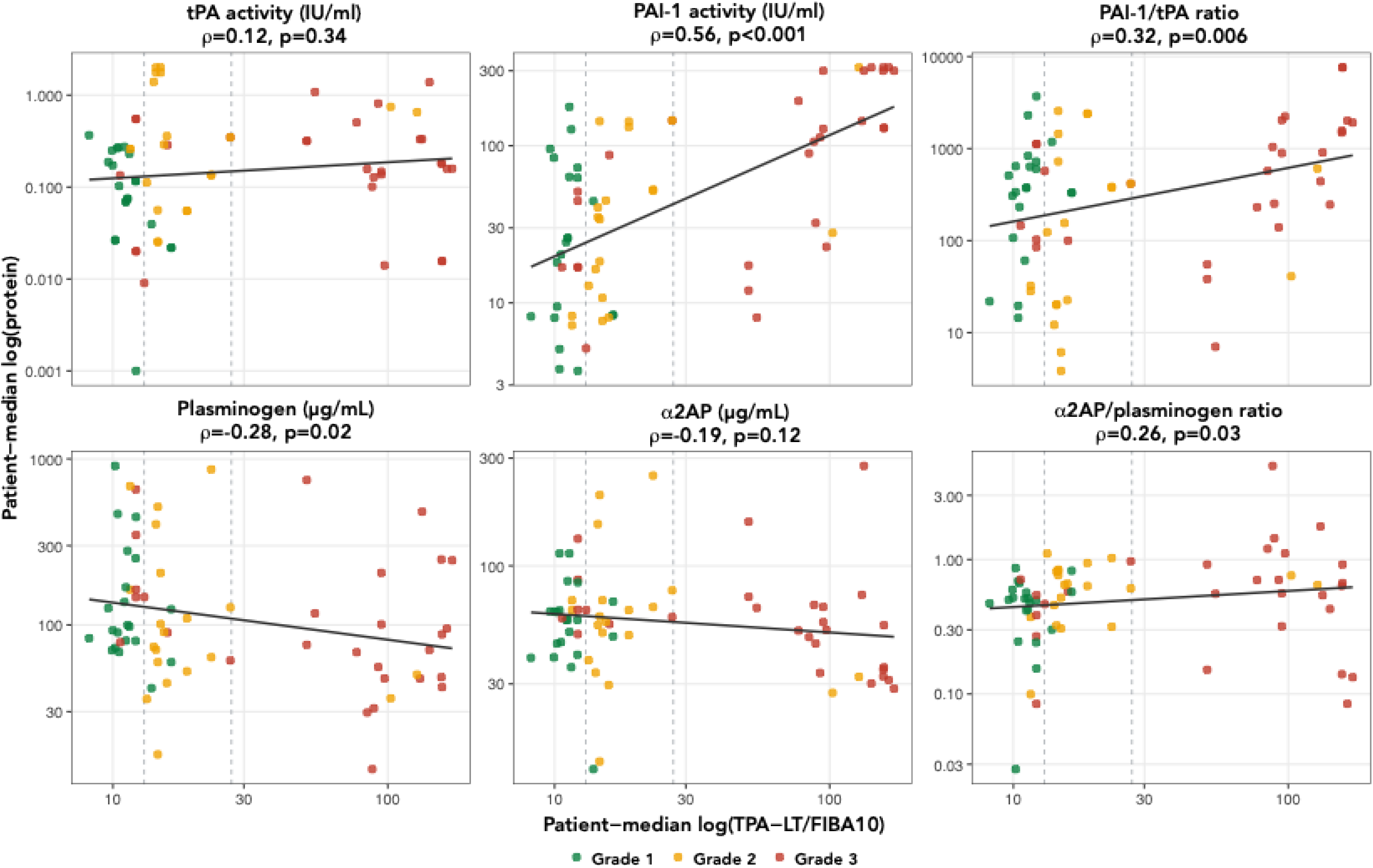
Patient level linear log-log correlations between fibrinolysis proteins and TPA-LT/FIBA10. Each panel reports the Spearman correlation coefficient (ρ) with its p value

**Figure S5.**
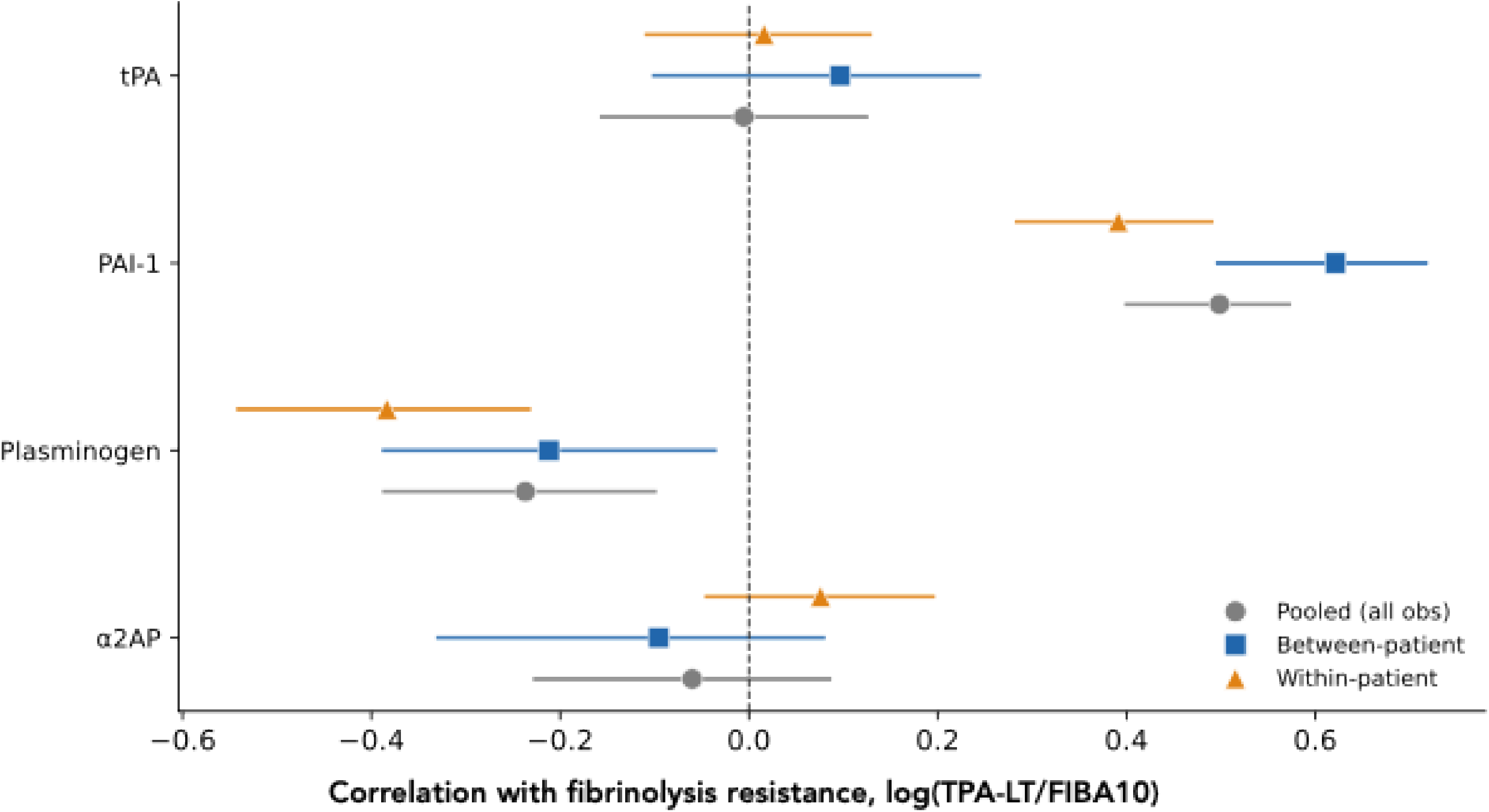
Within-patient and between-patient comparisons of correlations between fibrinolysis proteins and VET-derived TPA-LT/FIBA10. Points are estimates, bars are patient-bootstrapped 95% confidence intervals. A positive value represents an increase and a negative value represents a decrease.

**Figure S6.**
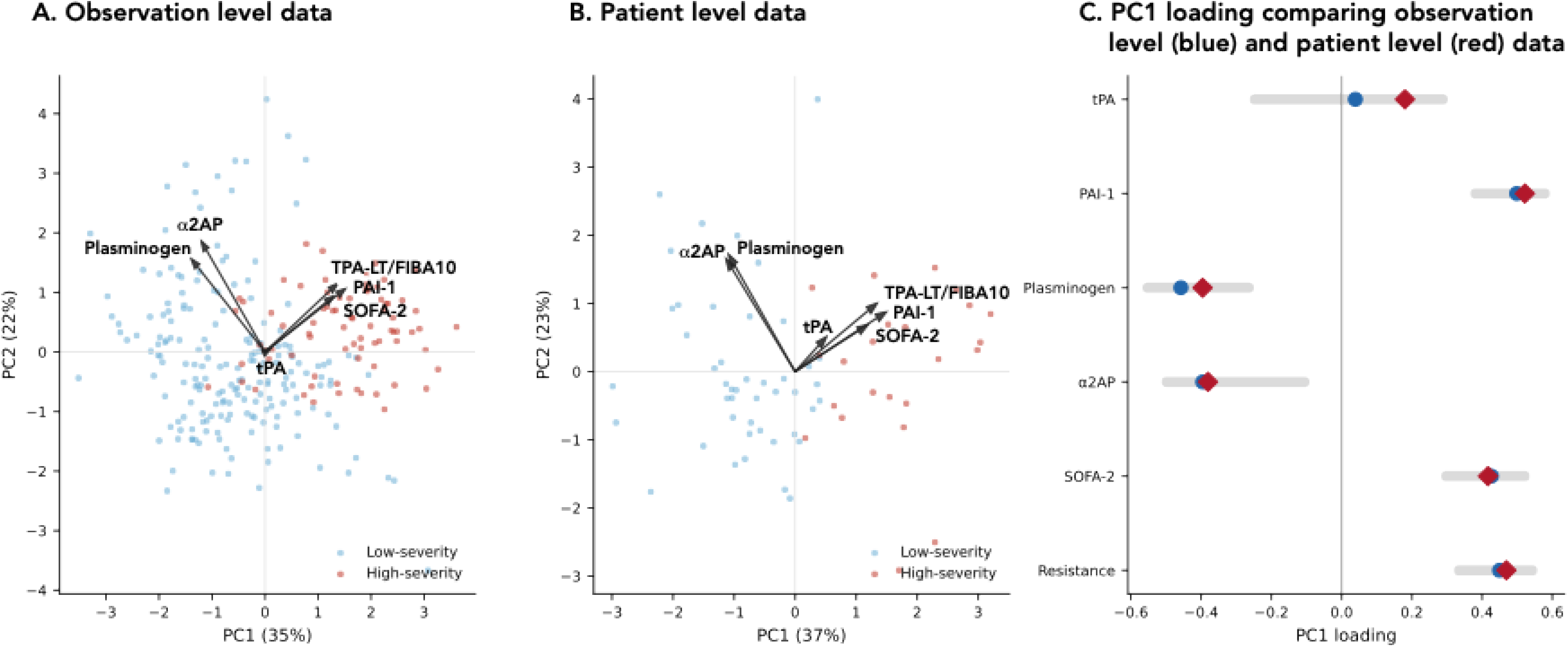
Principal component analysis (PCA) showing loadings using observation (panel A) and patient level (panel B) data. Comparisons in PC1 loadings using patient level data (red diamonds) and observation level data (blue circles) including the 95% confidence intervals are shown in panel C. Note that patient level data showed the same PC1 and PC2 loadings with a slight increase in the joint variability (37% vs 35%).

